# Race and ethnicity in the landmark hypertrophic cardiomyopathy trials: representation against population, site-catchment, and disease-referenced expectations

**DOI:** 10.64898/2026.09.24.26363977

**Authors:** Andrew P Wescott, Bradley J Petek, Jack W. O’Sullivan, Farid Moussavi-Harami, David Owens, Allan Lawrie, Christopher M. Kramer, Florian Rader, Victoria N Parikh, Matthew T Wheeler, Eugene Yang, Euan A Ashley, Ahmad Masri, Daniel Seung Kim

## Abstract

**BACKGROUND:** Hypertrophic cardiomyopathy (HCM) is a pan-ethnic disease, at least as common in Black as in White Americans, yet Black patients reach specialist care later, undergo septal reduction less often, and die sooner. Whether the defining trials enrolled representatively is unknown. We sought to measure Black participation in the landmark HCM trials against three expectations of increasing stringency, and to test whether trial site geography or quantitative eligibility thresholds account for the shortfall.

**METHODS:** Of 197 registered interventional HCM trials, 32 met prespecified landmark tiers; demographics were extracted for 16 and 13 reported race. The Black share of Black plus White participants was pooled by random-effects meta-analysis and compared with the US adult population, with each trial’s own site counties, and with a disease-referenced echocardiographic expectation. Site geography and eligibility thresholds were tested using site addresses, census data, and NHANES.

**RESULTS:** Across 13 trials, 70 of 1,780 Black or White participants were Black. The pooled Black share was 3.98% (95% CI 2.68-5.86), against a population expectation of 16.3%, giving a participation-to-prevalence ratio (PPR) of 0.24 (95% CI 0.16-0.36), the lowest of any racial and/or ethnic minority group. The clinical trial site network reached 63.7% of Black and 47.5% of White US adults, with geospatial analysis predicting 26.4% Black enrollment; however, every trial fell below this expectation. Quantitative eligibility thresholds closed only 2.0% of the enrollment gap.

**CONCLUSIONS:** Black participation was about one quarter of the population expectation. Neither site geography nor eligibility thresholds explain it.

**CLINICAL PERSPECTIVE:** *What is New?:*

- Across the 13 landmark hypertrophic cardiomyopathy trials that posted race, 70 of 1,780 participants recorded as Black or White were Black, a pooled share of 3.98% against a population expectation of 16.3%.
- The US counties hosting the trials’ own sites predicted 26.4% Black enrollment, and every trial enrolled below its own catchment expectation.
- Neither site geography nor quantitative eligibility thresholds explain the shortfall, because the site network reached 63.7% of Black adults and the extracted thresholds closed 2.0% of the gap in Black enrollment.

*What are the Clinical Implications?:*

- Black patients with hypertrophic cardiomyopathy live within reach of the sites that enrolled them least, so clinicians should treat trial referral as a distinct clinical act and audit their own referral patterns.
- Future trials should report screening, referral, and offer by race prospectively, and should benchmark enrollment against the catchment population of their own sites.
- The loss occurs downstream of geography and eligibility, in site recruitment conduct and in the referral pathway, and that is where measurement and correction must begin.

## INTRODUCTION

Hypertrophic cardiomyopathy (HCM) is the most common inherited cardiac disease, with a phenotypic prevalence near 1 in 500 adults^1^. Two guideline revisions in five years have reshaped the management of HCM^2,3^, and cardiac myosin inhibitors have moved from first-in-human study to FDA approval for obstructive HCM in under a decade^4,5^.

HCM does not spare Black Americans. A population echocardiographic survey of 4,111 young adults in the Coronary Artery Risk Development in Young Adults (CARDIA) study found unexplained left ventricular hypertrophy about 2.4 times more often in Black than in White participants^6^. Yet Black patients reach specialist care later, undergo septal reduction therapy less often, and have higher rates of heart failure and death^7,89^. Similar patterns appear in Afro-Carribean cohorts^10^. Sudden death in young Black athletes is disproportionately attributed to HCM^11,12^. Variant interpretation has identified many pathogenic mutations in Black populations^13^, and registry data show that genotype and phenotype together drive lifetime burden^14^. The disease is therefore not rare in Black Americans, and although HCM-related mortality halved nationally between 1999 and 2019, it remained higher in Black adults throughout^15^.

Racial and ethnic representation in trials has been measured, demonstrating that Black patients are consistently under-enrolled relative to disease burden^16,17^. Reporting standards and federal policy have tightened in response: journals now require explicit race and ethnicity reporting^18^, the Office of Management and Budget revised its statistical standards in 2024^19^, and the Food and Drug Administration has issued draft guidance requiring sponsors to file Diversity Action Plans^20^.

Measurement has also improved alongside policy. Enrollment can now be compared with the intended-use population, with the catchment areas of the sites actually recruited, or with disease-referenced prevalence^21–23^. Eligibility criteria are a separate and testable mechanism, because thresholds set on laboratory values can exclude groups differentially (we use ceiling for an upper threshold limit and floor for a lower threshold limit throughout). Natriuretic peptide floors are the clearest example, since Black patients carry systematically lower levels at equal disease severity^24^. Kidney function floors have their own history, and race-based estimating equations were retired only recently^25,26^.

In the current study, we assembled the landmark trials in HCM and measured Black participant representation against three benchmarks of increasing stringency: the United States (US) adult population, the catchment areas of the sites that actually enrolled, and the disease-referenced prevalence. We then examined the two mechanisms most often invoked to explain such shortfalls: the geographic placement of trial sites and the quantitative eligibility thresholds written into protocols, to establish how much of the observed deficit in Black enrollment either process can account for.

## METHODS

### Study design and ethics

This is a cross-sectional analysis of the landmark HCM clinical trials. Trial-level participant demographics reported to a public trial registry were compared with three external benchmarks: the US adult population, the population of the geographic areas served by the enrolling sites, and disease-referenced prevalence. The site-catchment and eligibility-threshold analyses were undertaken as mechanistic follow-ups to the primary comparison and are reported as explanatory rather than confirmatory. Reporting follows STROBE recommendations for cross-sectional studies^27^.

All data sources are publicly available and contain only aggregate, de-identified information. Hence, consent for participation was not applicable to this work. The research was reviewed by the University of Washington Institutional Review Board and deemed not to meet definitions for human subjects research.

### Data sources

Trial characteristics, participant demographics, registered facility addresses, and verbatim eligibility criteria were obtained from the ClinicalTrials.gov version 2 application programming interface^28^, queried on September 1, 2026. Population denominators were taken from the American Community Survey (ACS) 2019 to 2023 five-year estimates (table B03002; Hispanic or Latino origin by race) at the county, place, core-based statistical area, and state level^29^. Non-Hispanic single-race counts were used throughout. The prevalence of quantitative eligibility criteria in the general adult population was estimated from the National Health and Nutrition Examination Survey (NHANES) 2017 to March 2020 pre-pandemic cycle^30^, supplemented for N-terminal pro-B-type natriuretic peptide (NT-proBNP) by the NHANES 1999 to 2004 surplus-sera assay file. The disease-referenced benchmark was taken from the published CARDIA echocardiographic survey of unexplained left ventricular hypertrophy in young adults^6^.

### Trial identification and selection

We queried interventional studies of HCM with no restriction on recruitment status, phase, sponsor, or date, which returned 197 records. Studies were eligible for screening if they were phase 2, 3, or 4, and enrolled participants with HCM. Phase 1, early-phase 1, and combined phase 1/2 studies were excluded as uninformative about the populations exposed to therapies in late-stage development. Similarly, studies carrying no registered phase (which were predominantly investigator-initiated procedural, device, and/or diagnostic studies) were excluded. Studies without a registered phase were likewise excluded, since these were predominately investigator-initiated procedural, device, and diagnostic studies. Studies enrolling for another cardiomyopathy or heart failure phenotype (e.g., ischemic or non-ischemic cardiomyopathy, Danon disease, Friedreich ataxia cardiomyopathy, and heart failure with preserved ejection fraction) were excluded. One study without a registered phase was retained as phase labeling is not applied to behavioral interventions; this applied to one exercise training trial in HCM (RESET-HCM, NCT01127061)^31^.

### Landmark classification

Three landmark tiers were defined, and tier assignment was completed before any demographic data were extracted. Tier 1 comprised trials cited in the clinical studies section of a current US product label for an HCM indication. Tier 2 comprised other sponsor-run registrational trials of an agent developed for HCM, including phase 2 dose-finding studies and open-label extensions, identified from the registry sponsor and intervention fields. Tier 3 comprised trials cited in the reference list of the 2020 or 2024 ACC/AHA HCM guidelines^3,32^. A trial satisfying more than one definition was assigned to the lowest-numbered tier it satisfied. Thirty-two (32) of the 83 screened studies met one or more tier definitions, comprising 4 label-pivotal, 24 registrational, and 4 guideline-cited trials.

### Analytic set

Of the 32 landmark trials, 19 had a results source, defined as a posted ClinicalTrials.gov results module or a retrievable primary publication reporting participant characteristics (a design or protocol paper alone did not qualify). Demographics were extracted for 16. The three that were not extracted were: INHERIT^33^ (enrolled only in Denmark and hence contributes no US race data) and two open-label extensions: mavacamten (NCT03723655)^34^ and aficamten (FOREST-HCM)^35^, which re-enroll participants from trials already in the analysis set. Thirteen of the 16 extracted trials reported race data in a form that separates the categories required for the primary comparison, and these 13 constitute the analytic set for representation.

### Demographic extraction

For each extracted trial, the posted results module was read and participant counts recorded for every race and ethnicity category together with the verbatim category labels. We recorded whether the five race categories were reported separately, whether ethnicity was reported separately from race, and the proportion of participants with unknown or unreported race. Where a trial reported Asian and Pacific Islander participants together, or combined race and ethnicity within a single combined schema, the record was flagged as not separable and excluded from the affected comparison rather than from the study. Published reports were used only to confirm registry values and never to replace them. Race and ethnicity are as recorded by investigators at enrolling sites, are predominantly self-identified, and are treated throughout as social rather than biologic constructs.

### Primary comparison and benchmarks

Registry race categories are Hispanic/Latinx-inclusive, whereas census population denominators separate Hispanic/Latinx ethnicity. Hence, a Hispanic-inclusive numerator over a non-Hispanic denominator distorts the ratio for White, Black, and Asian participants. To reduce the mismatch, the primary comparison was restricted to participants recorded as Black or White, and expressed as the Black share of that two-group total, following convention in US vital statistics of limiting race-specific comparisons to the two groups whose classification is concordant across numerator and denominator^36^. Bridging the denominator instead would require individual-level joint distribution of race and Hispanic origin, which registry-posted aggregate counts do not provide^37^. The corresponding population benchmark is the Black share of the Black plus White US adult population aged 20 years and over, which is 16.3% in ACS 2019-2023, computed from tables B01001B and B01001H29. The disease-referenced benchmark is the Black and White composition of participants with unexplained left ventricular hypertrophy in CARDIA, constructed by applying the race-specific prevalence ratio to the same population base, yielding 31.9% (note that this analysis is indicative rather than definitive, because it rests on a young adult cohort and a small number of affected participants)^6^.

The primary endpoint was the participation-to-prevalence ratio (PPR) for Black participants against the population benchmark, defined as the pooled trial share divided by the benchmark share, where 1.0 denotes proportional representation and values below 1.0 denote under-representation^22^. Secondary endpoints were the PPRs against the site-catchment and disease-referenced benchmarks, the observed-to-expected ratio within each trial, the completeness of race and ethnicity reporting, the geographic coverage of the Black adult population by the trial site network, and the differential exclusion of Black relative to White adults by quantitative eligibility thresholds.

We performed several denominator-sensitivity analyses, in which the Black share of the Black plus White HCM population was drawn from three contemporary sources that measure at different points of the care pathway: (1) hospitalized HCM in the National Inpatient Sample^8^, (2) diagnosed obstructive HCM in the Optum Market Clarity integrated clinical and claims database^38^, and (3) patients reaching an HCM specialty center in the Sarcomeric Human Cardiomyopathy Registry (SHaRe)^39,40^. These sources estimate the diagnosed and referred population, rather than the disease prevalence, and are therefore conservative by construction. The National Inpatient Sample and Optum exclude Hispanic patients from their Black and White counts, as the primary ACS benchmark does, so they introduce no mismatch beyond the one described above. SHaRe asks about race only, so its counts match the Hispanic-inclusive trial numerator directly.

### Site catchment

For the 13 analyzable trials with race data, every registered facility was extracted, yielding 654 site records, of which 281 were within the US. Each site was geocoded to a county and to a core-based statistical area by matching the registered city and state against census place and county subdivisions^41^. For each trial, the expected Black share is the unweighted mean of the county-level Black shares of the Black plus White population across that trial’s US site records, which weights a trial’s counties by how many of its sites sit in them; the observed-to-expected ratio divides the trial’s observed Black share by this catchment expectation^42,43^. The pooled catchment expectation of 26.4% is the population-weighted Black share across the unique counties hosting any of the 13 trials’ sites, and is the quantity the eligibility cascade is applied to^44^.

The same per-trial calculation was repeated at the metropolitan, place, and state level as sensitivity analyses. For the institution-level analysis, addresses were pooled across the 23 landmark trials; records carrying anonymized names such as “Research Site” or “Local Institution” were excluded from institution counts while being retained for geographic counts wherever city and state were present.

### Eligibility thresholds

Quantitative eligibility thresholds from the criteria text of all 32 landmark trials were extracted, recording body mass index (BMI) ceilings, hemoglobin floors, estimated glomerular filtration rate floors, creatinine ceilings, natriuretic peptide floors, age limits, and functional testing requirements. For each threshold, the fraction of adults excluded was estimated by race in NHANES using survey-weighted proportions^30,45^. Because NT-proBNP was not measured in the pre-pandemic cycle, the natriuretic peptide floor was evaluated in the 1999 to 2004 surplus-sera file^46^, and a linear model of log NT-proBNP on race, age, sex, and BMI among adults aged 20 years and over was fitted to obtain an adjusted geometric mean ratio. Thresholds were then applied sequentially to the pooled site-catchment expectation, each weighted by the share of the analyzed participant pool contributed by the trials that applied it. This cascade is an arithmetic upper bound on the effect of thresholds on the eligible pool, not a causal decomposition of enrollment.

### Statistical analysis

Analyses were performed using Python 3.11 with statsmodels, scipy, pandas, and geopandas.

Trial-level proportions were pooled by random-effects meta-analysis on the logit scale, using the Paule-Mandel estimator of between-trial variance^47^. Between-trial heterogeneity is reported as I^2^ ^48,49^. Trial-level proportion intervals are Clopper-Pearson exact intervals^50^. A continuity correction of 0.5 was applied only to trials with a zero cell, and the crude participant-weighted percentage is reported alongside the model estimate so that sparse strata can be recognized; strata in which nearly all cells are empty are not interpreted^51^. Confidence intervals for the population and site-catchment PPRs were propagated from the pooled estimate with the benchmark treated as fixed. The association between catchment expectation and observed enrollment was tested by Spearman rank correlation. Missing data were not imputed: trials without a results source, without race data, or without separable race and ethnicity categories were excluded from the affected comparison and the number of contributing trials is reported for every estimate. Tests were two-sided at the *α*=0.05 level, no adjustment was made for multiplicity, and the study is descriptive. Confidence intervals rather than P-values are the basis for inference throughout.

### Data and code availability

All source data are public. The extracted trial-level tables, benchmark tables, and analysis code are provided via permanent digital object identifier (10.5281/zenodo.22926460).

## RESULTS

### Trials and reporting

Of 197 registered interventional HCM trials, 83 met screening criteria, 32 met a landmark definition, and 19 had a results source (**Figure S1**). Demographics were extracted for 16, comprising 3 label-pivotal, 10 registrational, and 3 guideline-cited trials. Thirteen trials reported race data, covering 2,126 enrolled participants (**Table 1**). Race reporting was complete in all 3 label-pivotal trials, in 8 of 10 registrational trials, and in 2 of 3 guideline-cited trials, so 3 of the 16 extracted landmark trials reported no race data at all (**Table S1**).

**Table 1.** Characteristics of the 13 landmark HCM trials with posted race data.

| **Trial** | **NCT** | **Phase** | **Intervention** | **Sponsor** | **Start** | **Enrolled (N)** | **US Sites (N)** | **Black (N)** | **White (N)** | **Black+White (N)** | **Black (%)** | **Evidence tier** |
| --- | --- | --- | --- | --- | --- | --- | --- | --- | --- | --- | --- | --- |
| RESET-HCM | NCT01127061 | N/A | Exercise training | Academic | 2010 | 136 | 2 | 5 | 119 | 124 | 4.0% | Guideline-cited |
| VANISH | NCT01912534 | 2 | Valsartan | Academic | 2014 | 211 | 13 | 3 | 173 | 176 | 1.7% | Guideline-cited |
| EXPLORER-HCM | NCT03470545 | 3 | Mavacamten | Industry | 2018 | 251 | 29 | 6 | 229 | 235 | 2.6% | Label-pivotal |
| SEQUOIA-HCM | NCT05186818 | 3 | Aficamten | Industry | 2022 | 282 | 48 | 3 | 223 | 226 | 1.3% | Label-pivotal |
| VALOR-HCM | NCT04349072 | 3 | Mavacamten | Industry | 2020 | 112 | 21 | 3 | 100 | 103 | 2.9% | Label-pivotal |
| LIBERTY-HCM | NCT02291237 | 2/3 | Eleclazine | Industry | 2015 | 172 | 26 | 10 | 147 | 157 | 6.4% | Registrational |
| MAPLE-HCM | NCT05767346 | 3 | Aficamten | Industry | 2023 | 175 | 28 | 1 | 140 | 141 | 0.71% | Registrational |
| MAVERICK-HCM | NCT03442764 | 2 | Mavacamten | Industry | 2018 | 59 | 32 | 2 | 52 | 54 | 3.7% | Registrational |
| MERCUTIO | NCT05556343 | 2 | MYK-224 | Industry | 2023 | 18 | 12 | 0 | 17 | 17 | 0% | Registrational |
| ODYSSEY-HCM | NCT05582395 | 3 | Mavacamten | Industry | 2022 | 580 | 41 | 25 | 400 | 425 | 5.9% | Registrational |
| PIONEER-HCM | NCT02842242 | 2 | Mavacamten | Industry | 2016 | 21 | 7 | 1 | 20 | 21 | 4.8% | Registrational |
| PIONEER-OLE | NCT03496168 | 2 | Mavacamten | Industry | 2018 | 13 | 4 | 1 | 12 | 13 | 7.7% | Registrational |
| REDWOOD-HCM | NCT04219826 | 2 | Aficamten | Industry | 2020 | 96 | 18 | 10 | 78 | 88 | 11.4% | Registrational |

### Representation

Among the 2,126 participants enrolled in landmark HCM trials, 1,780 (83.7%) were recorded as Black or White, of whom 70 were Black. The pooled Black share was 3.98% (95% CI 2.68-5.86), with moderate heterogeneity (I^2^ 53.7%). Against the population expectation of 16.3%, the PPR was 0.24 (95% CI 0.16-0.36). Against the disease-referenced CARDIA expectation of 31.9%, the PPR was 0.12 (95% CI 0.08-0.18). The deficit did not depend on the benchmark. Against the diagnosed HCM population, the PPR was 0.19 (95% CI 0.13-0.28) using Optum^38^ and 0.18 (95% CI 0.12-0.27) using the National Inpatient Sample^8^, and against the specialty referral-center population in SHaRe^14,39^ it was 0.39 (95% CI 0.27-0.58; **Table S2**).

On the all-race denominator (which is race/ethnicity inclusive and is the like-for-like comparison across groups), the Black PPR was 0.30 (95% CI 0.20-0.45), the lowest of any interpretable group. Hispanic/Latinx participants were also under-represented at 0.44 (95% CI 0.28-0.67), and the Asian PPR was 0.79 (95% CI 0.45-1.35), possibly inflated by the Hispanic-inclusive numerator. American Indian, Alaska Native, Native Hawaiian, and Pacific Islander strata contained no or almost no participants, and their pooled values reflect the continuity correction rather than data (**Table 2**, **Figure 1**, **Figure 2**).

**Figure 1.**
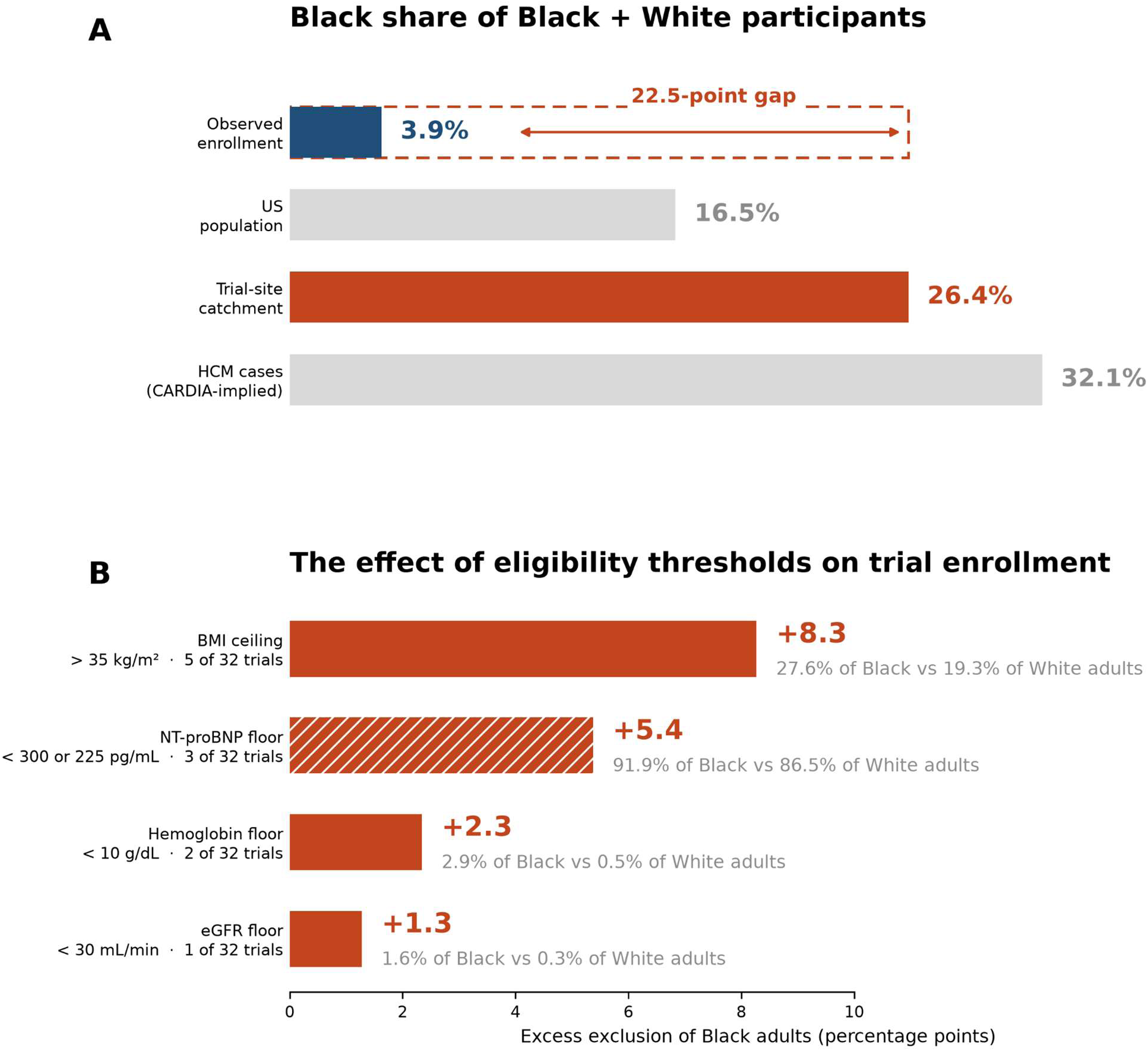
Black participation in the landmark HCM trials, and what does not explain it. (A) Black share of Black plus White participants across the 13 landmark trials with posted race data, against three expectations: the US adult population, the population-weighted composition of the counties hosting the trials’ own sites, and unexplained left ventricular hypertrophy in CARDIA. The dashed outline marks the 22.5-point gap to the site-catchment expectation. (B) Excess exclusion of Black over White adults under each eligibility threshold, in percentage points, with the underlying exclusion percentages and the number of the 32 trials applying each threshold. Prevalences are from NHANES 2017-2020, expected from the hatched NT-proBNP bar (NHANES 1999-2004). Applied in cascade, these thresholds explain 0.46 of the 22.5-point gap. Navy, observed enrollment. Red - the site-catchment expectation and disparities derived from it; Gray - contextual benchmarks.

**Figure 2.**
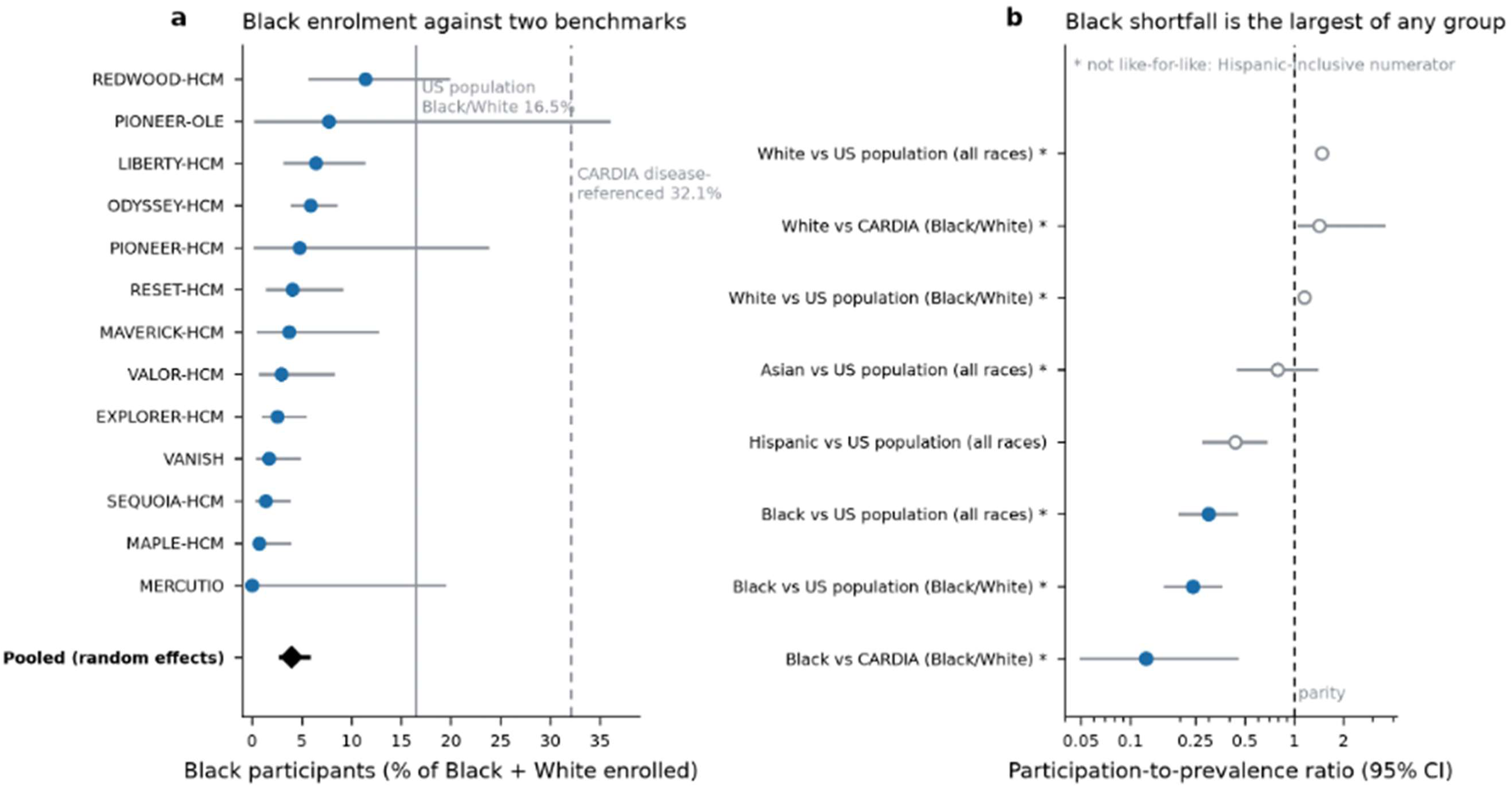
Representation against benchmarks, and participation-to-prevalence ratios by group. (A) Black share of Black plus White participants in each of the 13 landmark trials, with Clopper-Pearson exact intervals and random-effects pooled estimates (black diamond). Reference lines mark the population (solid) and CARDIA disease-referenced (dashed) expectations. (B) Participation-to-prevalence ratios with 95% confidence intervals against the US adult population; values below 1.0 indicate under-representation. The Asian and White ratios are not like-for-like (*), because the registry numerator is Hispanic-inclusive, while the census denominator is not. Strata with all or nearly all cells zero are omitted.

**Table 2.** Pooled racial and ethnic representation against population and disease-referenced benchmarks. Crude (%) is the participant-weighted aggregate across eligible HCM trials. Pooled % is the random-effects estimate on the logit-scale (using Paule-Mandel between-trial variance), which weights trials by inverse variance rather than by size, therefore estimating the share in a typical trial rather than in the assembled participant pool. The pooled value is not interpretable when nearly all cells are zero (as is the case for *AIAN and NHPI).

| Racial and/or Ethnic Group | Trials (N)** | Total Participants (N) | Crude (N) | Crude % | Pooled % (95% CI) | Benchmark (%) | PPR (95% CI) |
| --- | --- | --- | --- | --- | --- | --- | --- |
| *American Community Survey 2019-2023 (Table B03002), Population Share of Age 20+* |  |  |  |  |  |  |  |
| White | 13 | 2092 | 1710 | 81.7 | 86.8 (81.7-90.7) | 59.2 | 1.47 (1.38-1.53) |
| Black | 13 | 2092 | 70 | 3.35 | 3.51 (2.35-5.21) | 11.7 | 0.30 (0.20-0.45) |
| Asian | 12 | 1914 | 193 | 10.1 | 5.53 (3.16-9.51) | 6.98 | 0.79 (0.45-1.35) |
| AIAN* | 12 | 1914 | 4 | 0.21 | 0.66 (0.33-1.31)* | 0.66 | 1.00 (0.50-1.99) |
| NHPI* | 11 | 1742 | 0 | 0 | 0.54 (0.23-1.25)* | 0.19 | 2.81 (1.22-6.46) |
| Hispanic | 12 | 1810 | 188 | 10.4 | 8.05 (5.13-12.4) | 18.5 | 0.44 (0.28-0.67) |
| *American Community Survey 2019-2023 (Tables B0100B and B01001H), Population Share of Age 20+, Black/White restricted denominator* |  |  |  |  |  |  |  |
| White | 13 | 1780 | 1710 | 96.1 | 96.0 (94.1-97.3) | 83.7 | 1.15 (1.12-1.16) |
| Black | 13 | 1780 | 70 | 3.93 | 3.98 (2.68-5.86) | 16.3 | 0.24 (0.16-0.36) |
| *CARDIA Disease Prevalence, Black/White restricted denominator* |  |  |  |  |  |  |  |
| White | 13 | 1780 | 1710 | 96.1 | 96.0 (94.1-97.3) | 68.1 | 1.41 (1.38-1.43) |
| Black | 13 | 1780 | 70 | 3.93 | 3.98 (2.68-5.86) | 31.9 | 0.12 (0.08-0.18) |

Abbreviations: AIAN - American Indian and Alaskan Native; NHPI - Native Hawaiian and Pacific Islander; PPR - participation-to-prevalence ratio
\*Note that data for AIAN and NHPI participants was extremely sparse. Nearly all cells were zero. The pooled value is driven by the continuity correction and should be interpreted with caution.
\*\*Only trials that contributed data are counted. Hence, if a trial reported no row for a category (e.g., VANISH reported no Asian, AIAN, NHPI participants; LIBERTY-HCM reported no NHPI counts), they are not counted in this column.

The deficit was stable across analysis sets (**Table S3**). The pooled Black share ranged from 2.24% (95% CI 1.28-3.90) in the 3 label-pivotal trials, to 6.51% (95% CI 3.82-10.89) in the 7 US-only trials of any tier. It was 5.98% (95% CI 3.87-9.12) in the registrational tier and 3.92% (95% CI 2.24-6.77) in the 5 US-only landmark trials, a set that includes the exercise training trial RESET-HCM^52^ and the non-obstructive disease trial MAVERICK-HCM^53^. It was 3.58% (95% CI 1.91-6.61) in the 8 multinational landmark trials. No stratum reached half the population expectation.

### Site catchment

The 13 trials registered sites in counties whose Black plus White populations were 26.4% Black, a higher expectation than the national 16.3% because trial sites concentrate in metropolitan counties. Every trial enrolled below its own catchment expectation (**Table 3**, **Figure 3**). Observed-to-expected ratios ran from 0.00 in MERCUTIO, which enrolled 18 participants, to 0.41 in REDWOOD-HCM^54^. Twelve of 13 upper confidence intervals limits excluded parity, the exception being the 13-participant PIONEER open-label extension^55^. The three label-pivotal trials, EXPLORER-HCM^4^, VALOR-HCM^56^, and SEQUOIA-HCM^5^, had ratios of 0.11, 0.11, and 0.05, respectively. Defining catchment at the metropolitan, city, or state level rather than the county level left all 13 trials below parity, with the highest ratio at any definition reaching 0.76 (**Table S4**). Catchment expectation and observed enrollment were unrelated across trials (Spearman *ρ*=0.21, *P*=0.49), as were catchment expectation and the observed-to-expected ratio *ρ*=0.05 *P*=0.87).

**Figure 3.**
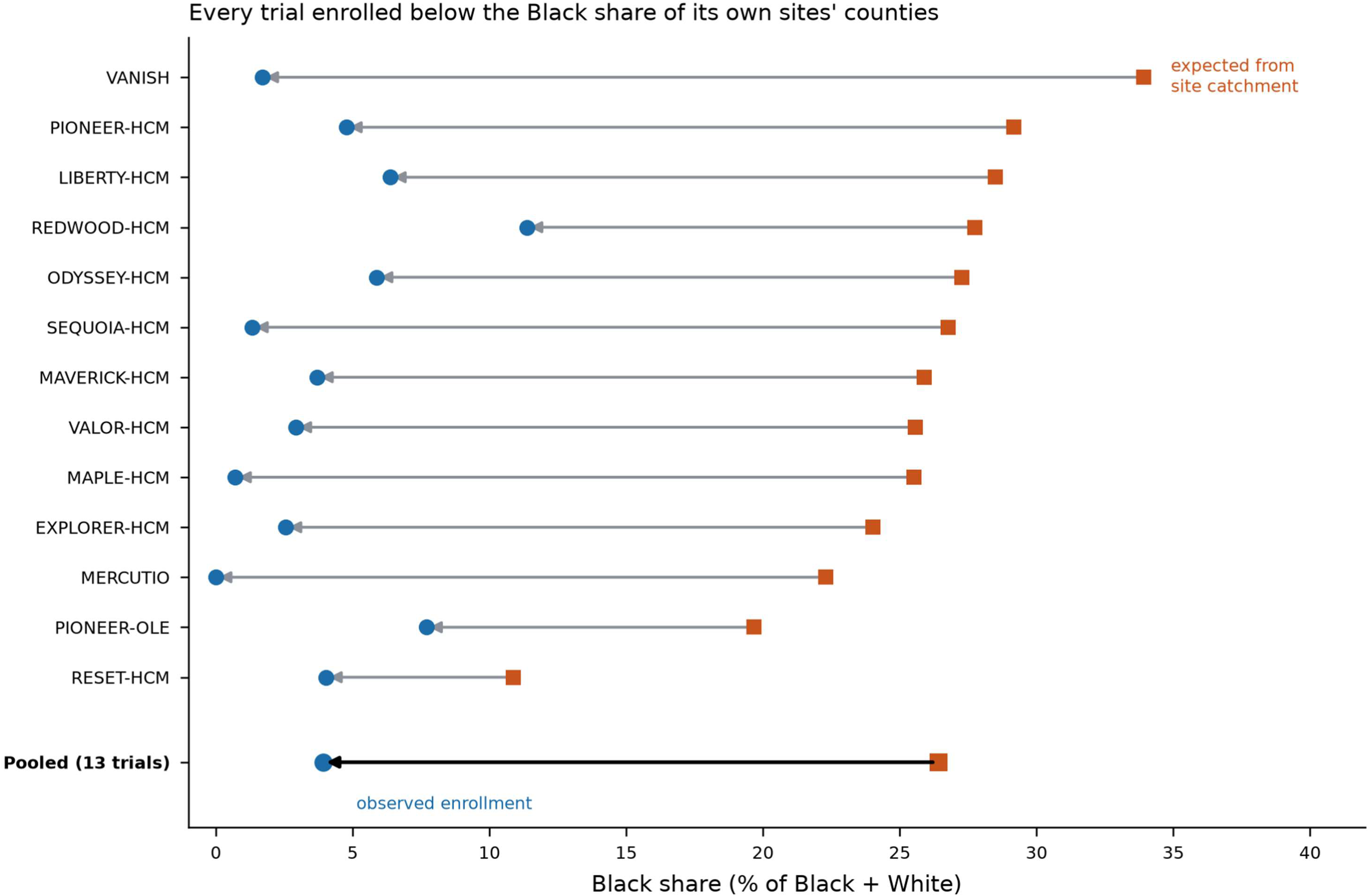
Every trial enrolled below its own site-catchment expectation. For each of the 13 landmark trials, the orange square marks the Black share expected from the counties hosting its registered sites and the circle marks the observed Black share, with an arrow from expectation to observation. Trials are ordered by expectation; ratios and intervals are in Table 3.

**Table 3.**
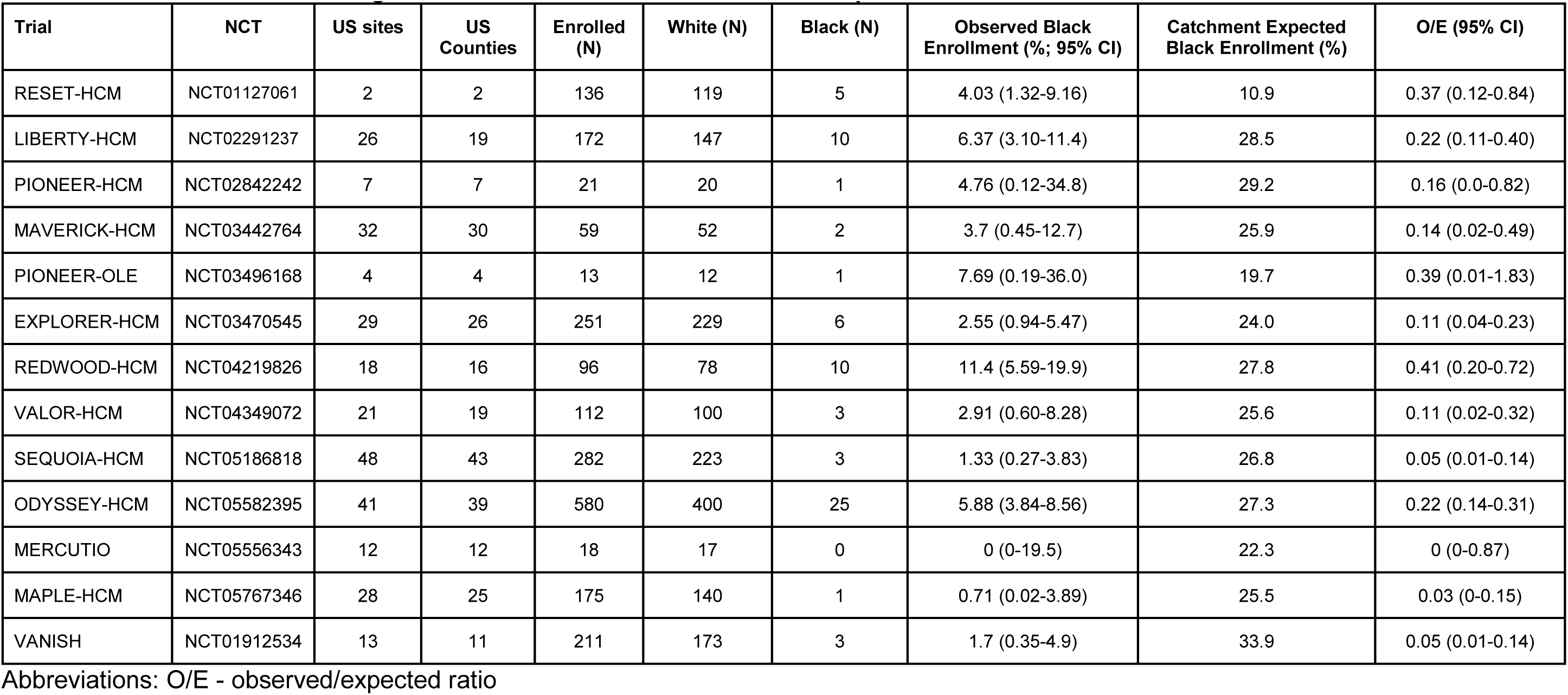
Observed enrollment against each trial’s own site-catchment expectation.

### Geography

Site placement did not disadvantage Black Americans. Counties hosting at least one trial site contained 39.2% of the Black non-Hispanic US population and 22.5% of the White non-Hispanic population. Metropolitan areas hosting a site contained 63.7% and 47.5% of the Black and White non-Hispanic US adult populations, respectively. The comparison survives restriction to metropolitan residents: among adults who live in any metropolitan area at all, 69.9% of Black residents and 58.0% of White residents lived in a metropolitan area hosting a trial site. Twenty-three of the 25 largest Black population metropolitan areas hosted a site, as did 25 of the 30 largest (**Figure 4**). Population coverage percentages are computed from complete census counts, rather than from a sample, and are therefore reported without confidence intervals.

**Figure 4.**
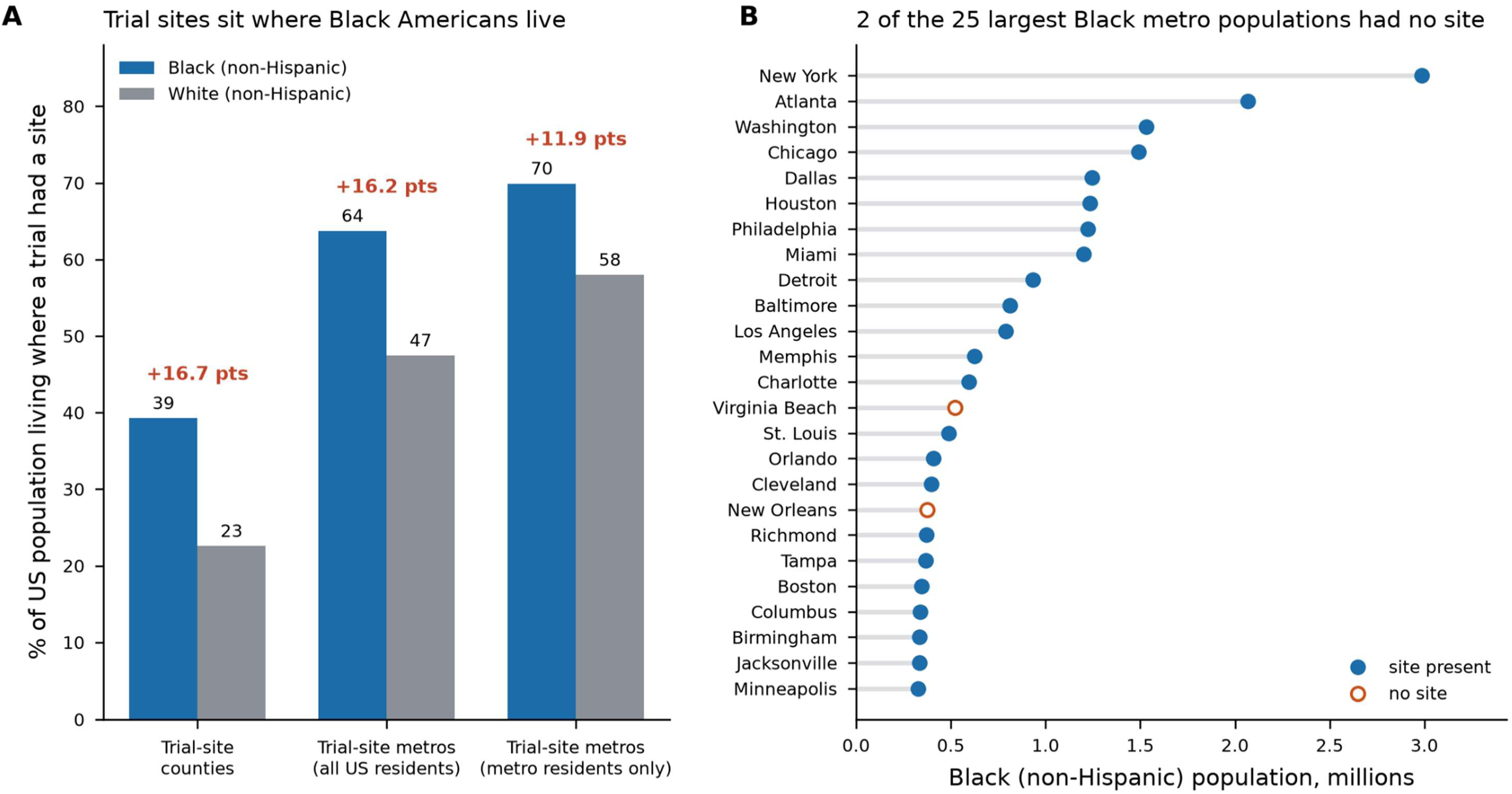
The trial site network reaches the Black population more than the White population. (A) Shares of the US Black and White non-Hispanic adult populations living where a landmark trial had a site, at three definitions: site-hosting counties, site-hosting metropolitan areas, and site-hosting metropolitan areas among metropolitan residents only.The first two are shares of the entire non-Hispanic Black and White populations, 40 million and 193.4 million. The third restricts both the numerator and denominator to the 36.4 million Black and 158.3 million White residents of metropolitan areas. Denominators are American Community Survey 2019-2023 five-year estimates, table B03002. The Black minus White difference in percentage points is given above each pair. (B) The 25 metropolitan areas with the largest Black adult population, in order, marking the two areas without an HCM trial site.

Trial networks were institutionally concentrated. Across the 23 landmark trials with registered US site addresses, 245 unique facilities appeared, of which 180 were named. Of those 180, 85 (47.2%) appeared in two or more landmark trials, and 26 (14.4%) appeared in five or more. One center appeared in 11 (**Table S5**).

### Eligibility thresholds

Of the thresholds extracted (**Table S6**), three of the 13 analyzable trials applied a BMI ceiling at any value (two of them at 35 kg/m^2^), and two applied a hemoglobin floor. No analyzable trial applied an estimated glomerular filtration rate floor. In NHANES, the fraction of adults above a BMI of 35 was 27.6% among Black adults and 19.3% among White adults, and the fraction with hemoglobin below 10 g/dl was 2.9% and 0.5%, respectively (**Table S7**).

Every threshold ran in the same direction, excluding a larger share of Black than White adults. The BMI ceiling of 35 kg/m^2^, applied by 5 of the 32 trials, had the largest absolute effect, excluding 27.6% of Black against 19.3% of White adults for an excess of 8.3 percentage points. The hemoglobin floor of 10 g/dL contributed 2.3 percentage points and the estimated glomerular filtration rate (eGFR) floor of 30 mL/min/1.73 m^2^ contributed 1.3 percentage points (**Table S8**, **Figure 1**). These are unweighted prevalence differences, and are not the same quantity as the threshold’s contribution to the eligibility cascade, which is zero for any criterion that no analyzable trial applied (as is the case for the eGFR floor, which was not applied in any of the 13 analyzable trials).

Applied in cascade to the site-catchment expectation, these thresholds moved it from 26.41% to 25.95%, closing 0.46 of the 22.48 percentage-point gap to observed Black enrollment, or 2.0% (**Table S9, Figure S2**). The cascade counts three analyzable trials at the BMI step, rather than the two that set the ceiling at 35 kg/m^2^, because PIONEER-HCM set its ceiling at 37 kg/m^2^. The remaining 22.02 points are unexplained by geography or threshold arithmetic (**Figure 1**). Even at universal application, meaning if every analyzable trial had applied all three extracted thresholds, the cascade could close no more than 12.1% of the Black enrollment gap.

The natriuretic peptide floor behaves differently from the anthropometric and laboratory ceilings, because a prevalence count does not capture its differential effect. Three of the 32 trials set a minimum NT-proBNP, two of them at 300 pg/mL and one at 225 pg/mL, and one of the trials at 300 pg/mL is in the analyzable pool. In the NHANES surplus-sera measurements, median NT-proBNP was 35.4 pg/mL in Black adults against 65.5 pg/mL in White adults, and 91.9% against 86.5% fell below 300 pg/mL (**Table S10**). Adjusted for age, sex, and BMI, the geometric mean NT-proBNP in Black adults was 0.71 times that in White adults (95% CI 0.67 to 0.75). A Black patient with HCM may therefore reach a fixed entry floor at a higher burden of disease than a White patient with the same physiology, and is more likely to be screened out at any given severity (**Figure 1**). Of note, two trials lowered the entry threshold for Black participants, an explicit attempt to correct for this distribution shift: ACACIA-HCM and a phase 2 study of EDG-7500 both set the entry threshold to 225 pg/mL for Black participants. However, neither has posted results, so neither contributes to our estimates.

## DISCUSSION

This cross-sectional analysis of the 32 landmark HCM trials shows that Black Americans received about one quarter of the clinical trial participation that their share of the adult US population would predict, and about one eighth of what a disease-referenced expectation would predict. The finding is not an artifact of one trial, one sponsor, or one era. It holds in the trials that supported the labels, in the trials that supported the guidelines, and in every sensitivity stratum we examined.

The two mechanisms most often invoked do not account for the lack of Black participation in clinical trials for HCM. First, trials are said to recruit where Black Americans do not live^57^. However, in HCM the opposite is true: the site network reached a larger share of the Black adult population than of the White, at both the county and metropolitan definition and also among the metropolitan residents alone. In addition, almost every large Black population metropolitan area hosted a site. This mirrors the catchment-based approach now used in cancer center accreditation^23^. Moreover, catchment expectation carried no association with enrollment across trials, so the sites that sat in the most heavily Black counties and cities did no better in recruiting Black Americans than the rest. Sponsors therefore placed sites appropriately where Black patients with HCM live, and Black enrollment in HCM trials fell short regardless, including against each trial’s own catchment. The shortfall is therefore not attributable to sponsor site selection.

The second mechanism is protocol arithmetic^58^. Eligibility criteria can exclude differentially, and in heart failure the natriuretic peptide floor is the recognized offender, since Black patients carry lower levels at equal severity^24^. In the landmark HCM trials, the applied quantitative thresholds were too sparsely used to matter. Only 3 of the 13 analyzable trials set a BMI ceiling, and none set a kidney function floor, a pattern consistent with the retirement of race-based estimating equations^26^. This eligibility cascade closed only 2% of the Black enrollment gap as applied in our data, and would account for no more than 12.1% even at theoretical universal application.

Three further mechanisms lie outside of our eligibility cascade, because they act through phenotype rather than through a laboratory cut-point. First, Black patients with HCM have lower resting and provoked left ventricular outflow tract gradients, less severe systolic anterior motion of the mitral valve, and more apical hypertrophy than White patients^59^. Hence, the outflow gradient entry criteria that define the obstructive HCM trials will exclude Black participants more often. However, no population data source records outflow gradients, so this cannot be quantified in the same way as a laboratory or BMI threshold. Second, 23 of the 32 trials set a left ventricular ejection fraction floor of 50-60%, which NHANES cannot evaluate; in the general population, the prevalence of reduced ejection fraction does not differ between Black and White adults^60^. Third, treatment-resistant hypertension is more common among Black adults in national survey data^61^, which could bear on both the eligibility for HCM trials and the attribution of hypertrophy to hypertension. We note that neither left ventricular ejection fraction or hypertension have been specifically characterized by race within HCM cohorts and both warrant future direct measurement.

Hence, the remaining possible mechanisms to account for the Black enrollment gap in HCM trials are not distinguishable from registry data alone. The first possible mechanism is site conduct, meaning whether a center hosting a trial screens and enrolls the Black patients already within its own catchment. This depends on where recruitment effort is spent, which clinics and clinicians within a health system are engaged, and how study staff are deployed across practices that serve different populations. The second possible mechanism is the patient pathway to an offer of enrollment, comprising recognition of HCM in patients presenting with symptoms and left ventricular hypertrophy, referral to the specialist centers that hold the trials, physician offers of participation, and patient acceptance into the trial. The denominator sensitivity analysis (**Table S2**) narrows this further. Patients reaching the HCM specialty centers contributing to SHaRe are 10.1% Black on a Black plus White base, against 3.98% in the trials these centers host, or a 2.5-fold shortfall that persists after conditioning on referral. Referral to specialist care is therefore unlikely to be the whole explanation, which places more of the remaining deficit on site conduct: screening, offer, and enrollment among Black patients a center is already seeing. We note that SHaRe’s contributing centers are not the same set as the 245 trial sites, so this is directional rather than an exact decomposition. Finally, direct financial barriers are an unlikely explanation for a deficit specific to Black enrollment. Study-related costs in these trials are sponsor-funded, and participation does not require the participant to carry insurance, although non-medical costs such as travel and time are not covered and remain a documented barrier^62^. However, in a nationally representative survey, cost-related considerations were reported as very influential to trial participation less often by Black than by White adults^63^.

Both of these potential mechanisms have evidence in the general clinical trials literature, but are invisible in registry data^17^. Referral is the step most open to *post-hoc* audit, because it is already recorded in the electronic health record. However, the late presentation and lower procedural rates documented in Black HCM patients suggests that the loss begins well before recruitment^7^. Diagnostic misattribution may compound this, both through variant misclassification^13^, and through the reading of left ventricular hypertrophy as caused by hypertension rather than from cardiomyopathy^40^.

Three practical implications follow. First, sponsors should benchmark against their own site catchments, not against the national population. The catchment expectation was 60% higher than the population expectation, so using the national benchmark will understate the deficit. Because site placement is not where the failure of Black HCM patient recruitment occurred, catchment benchmarking is diagnostic, rather than corrective. Second, the two trials that lowered the NT-proBNP threshold for Black HCM participants to allow more participation, show that sponsors can act on a known biologic difference without waiting for guidance. Third, reporting must be complete before it can be audited. Three of the 16 extracted landmark trials reported no race data at all, and across all 23 trials with extracted results, non-industry sponsors reported race in only 3 of 8 trials. Journal and registry requirements should be enforced at the same, strict standard^18^.

### Limitations

Several limitations of our study should be considered. First, Registry race categories are Hispanic-inclusive, so the two-group restriction we used is a correction, but not a cure. Our Black estimates remain conservative for this reason, with bounding the unobserved Hispanic/Latinx composition at both extremes placing our like-for-like Black American PPR between 0.09 and 0.27, which brackets our estimate and does not alter our conclusions. Second, the disease-referenced benchmark (CARDIA) rests on a single cohort of young adults with a small number of affected participants, and should be read as indicative and hypothesis-forming. We note that the denominator sensitivity analysis showed that the conclusions do not vary, as the PPR remained at or below 0.39 against three independent contemporary denominator sources (**Table S2**). Third, site addresses locate a facility rather than a catchment, and county composition is a coarse proxy for the population a center serves; facility anonymization also limited the institutional analysis to 180 of 245 unique sites. Fourth, we could not measure referral, screening or offer, because those data are not posted. Finally, the eligibility cascade analyses assume independence between thresholds and omits criteria we could not quantify (most importantly functional testing requirements, which were recorded for all 32 trials and applied by 4 of the 13 analyzable ones, but which have no population analogue in NHANES).

### Conclusions

In this cross-sectional analysis of the trials that have defined modern HCM therapy, Black participation was roughly one-quarter of the population expectation and one-eighth of a disease-referenced expectation. Neither the geography of the trial sites nor the quantitative eligibility thresholds explain the shortfall. The barrier lies downstream, in how sites recruit from the patients already within their reach and in the pathway that brings those patients to an offer, and this is where measurement and correction must now begin to balance representation in clinical trials moving forward.

## Data Availability

All source data are public. The extracted trial-level tables, benchmark tables, and analysis code are provided via permanent digital object identifier ().

## Nonstandard Abbreviations and Acronyms

ACS: American Community Survey
CARDIA: Coronary Artery Risk Development in Young Adults
CI: confidence interval
HCM: hypertrophic cardiomyopathy
NHANES: National Health and Nutrition Examination Survey
NT-proBNP: N-terminal pro-B-type natriuretic peptide
PPR: participation-to-prevalence ratio

## REFERENCES

1. Semsarian C, Ingles J, Maron MS, Maron BJ. New Perspectives on the Prevalence of Hypertrophic Cardiomyopathy. J Am Coll Cardiol. 2015;65:1249–1254.

2. Members WC, Ommen SR, Mital S, Burke MA, Day SM, Deswal A, Elliott P, Evanovich LL, Hung J, Joglar JA, et al. 2020 AHA/ACC Guideline for the Diagnosis and Treatment of Patients With Hypertrophic Cardiomyopathy. Circulation. 2020;142:e558–e631.

3. Members WC, Ommen SR, Ho CY, Asif IM, Balaji S, Burke MA, Day SM, Dearani JA, Epps KC, Evanovich L, et al. 2024 AHA/ACC/AMSSM/HRS/PACES/SCMR Guideline for the Management of Hypertrophic Cardiomyopathy A Report of the American Heart Association/American College of Cardiology Joint Committee on Clinical Practice Guidelines. J. Am. Coll. Cardiol. 2024;

4. Olivotto I, Oreziak A, Barriales-Villa R, Abraham TP, Masri A, Garcia-Pavia P, Saberi S, Lakdawala NK, Wheeler MT, Owens A, et al. Mavacamten for treatment of symptomatic obstructive hypertrophic cardiomyopathy (EXPLORER-HCM): a randomised, double-blind, placebo-controlled, phase 3 trial. Lancet. 2020;396:759–769.

5. Maron MS, Masri A, Nassif ME, Barriales-Villa R, Arad M, Cardim N, Choudhury L, Claggett B, Coats CJ, Düngen H-D, et al. Aficamten for Symptomatic Obstructive Hypertrophic Cardiomyopathy. N. Engl. J. Med. 2024;390:1849–1861.

6. Maron BJ, Gardin JM, Flack JM, Gidding SS, Kurosaki TT, Bild DE. Prevalence of Hypertrophic Cardiomyopathy in a General Population of Young Adults: Echocardiographic Analysis of 4111 Subjects in the CARDIA Study. Circulation. 1995;92:785–789.

7. Patlolla SH, Schaff HV, Nishimura RA, Eleid MF, Geske JB, Ommen SR. Impact of Race and Ethnicity on Use and Outcomes of Septal Reduction Therapies for Obstructive Hypertrophic Cardiomyopathy. J. Am. Hear. Assoc. 2022;12:e026661.

8. Johnson DY, Waken RJ, Fox DK, Hammond G, Maddox KEJ, Cresci S. Inequities in Treatments and Outcomes Among Patients Hospitalized With Hypertrophic Cardiomyopathy in the United States. J. Am. Hear. Assoc.: Cardiovasc. Cerebrovasc. Dis. 2023;12:e029930.

9. Ntusi NAB, Sliwa K. Associations of Race and Ethnicity With Presentation and Outcomes of Hypertrophic Cardiomyopathy JACC Focus Seminar 6/9. J. Am. Coll. Cardiol. 2021;78:2573– 2579.

10. Sheikh N, Papadakis M, Panoulas VF, Prakash K, Millar L, Adami P, Zaidi A, Gati S, Wilson M, Carr-White G, et al. Comparison of hypertrophic cardiomyopathy in Afro-Caribbean versus white patients in the UK. Heart. 2016;102:1797.

11. Petek BJ, Churchill TW, Moulson N, Kliethermes SA, Baggish AL, Drezner JA, Patel MR, Ackerman MJ, Kucera KL, Siebert DM, et al. Sudden Cardiac Death in National Collegiate Athletic Association Athletes: A 20-Year Study. Circulation. 2024;149:80–90.

12. Maron BJ, Haas TS, Ahluwalia A, Murphy CJ, Garberich RF. Demographics and Epidemiology of Sudden Deaths in Young Competitive Athletes: From the United States National Registry. Am. J. Med. 2016;129:1170–1177.

13. Manrai AK, Funke BH, Rehm HL, Olesen MS, Maron BA, Szolovits P, Margulies DM, Loscalzo J, Kohane IS. Genetic Misdiagnoses and the Potential for Health Disparities. N. Engl. J. Med. 2016;375:655–665.

14. Ho CY, Day SM, Ashley EA, Michels M, Pereira AC, Jacoby D, Cirino AL, Fox JC, Lakdawala NK, Ware JS, et al. Genotype and Lifetime Burden of Disease in Hypertrophic Cardiomyopathy. Circulation. 2018;138:1387–1398.

15. Minhas AMK, Wyand RA, Ariss RW, Nazir S, Khan MS, Jia X, Greene SJ, Fudim M, Wang A, Warraich HJ, et al. Demographic and Regional Trends of Hypertrophic Cardiomyopathy-Related Mortality in the United States, 1999 to 2019. Circ.: Hear. Fail. 2022;15:e009292.

16. Tahhan AS, Vaduganathan M, Greene SJ, Fonarow GC, Fiuzat M, Jessup M, Lindenfeld J, O’Connor CM, Butler J. Enrollment of Older Patients, Women, and Racial and Ethnic Minorities in Contemporary Heart Failure Clinical Trials: A Systematic Review. JAMA Cardiol. 2018;3:1011.

17. Michos ED, Reddy TK, Gulati M, Brewer LC, Bond RM, Velarde GP, Bailey AL, Echols MR, Nasser SA, Bays HE, et al. Improving the enrollment of women and racially/ethnically diverse populations in cardiovascular clinical trials: An ASPC practice statement. Am. J. Prev. Cardiol. 2021;8:100250.

18. Flanagin A, Frey T, Christiansen SL, Bauchner H. The Reporting of Race and Ethnicity in Medical and Science Journals. JAMA. 2021;325:1049–1052.

19. Federal Register:: Revisions to OMB’s Statistical Policy Directive No. 15: Standards for Maintaining, Collecting, and Presenting Federal Data on Race and Ethnicity [Internet]. [cited 2026 Sep 15];Available from: https://www.federalregister.gov/documents/2024/03/29/2024-06469/revisions-to-ombs-statistical-policy-directive-no-15-standards-for-maintaining-collecting-and

20. Diversity Action Plans to Improve Enrollment of Participants from Underrepresented Populations in Clinical Studies | FDA [Internet]. [cited 2026 Aug 31];Available from: https://www.fda.gov/regulatory-information/search-fda-guidance-documents/diversity-action-plans-improve-enrollment-participants-underrepresented-populations-clinical-studies

21. Sanyi A, Byiringiro S, Dabiri S, Jacobson M, Boyd A, Ogunniyi MO, Morris AA, Kohn R, Dickert NW, Lane-Fall MB, et al. Measuring Representativeness in Clinical Trials. Circulation. 2025;151:318–330.

22. Jenei K, Meyers DE, Prasad V. The Inclusion of Women in Global Oncology Drug Trials Over the Past 20 Years. JAMA Oncol. 2021;7:1569.

23. Rademacher N, Sisk C, Richman JS, Broman K, Wang C. Geospatial Analysis of Commission on Cancer-Accredited Centers Within Cancer Care Utilization–Based Catchment Areas. JCO Clin. Cancer Inform. 2025;9:e2500163.

24. Gangavelli A, Liu Z, Wang J, Okoh A, Steinberg R, Patel K, Pandey A, Gupta DK, Dickert N, Patel SA, et al. Racial differences in low natriuretic peptide levels: Implications for heart failure clinical trials. Am. Hear. J. 2023;265:1–10.

25. Delgado C, Baweja M, Crews DC, Eneanya ND, Gadegbeku CA, Inker LA, Mendu ML, Miller WG, Moxey-Mims MM, Roberts GV, et al. A Unifying Approach for GFR Estimation: Recommendations of the NKF-ASN Task Force on Reassessing the Inclusion of Race in Diagnosing Kidney Disease. Am. J. Kidney Dis. 2022;79:268–288.e1.

26. Inker LA, Eneanya ND, Coresh J, Tighiouart H, Wang D, Sang Y, Crews DC, Doria A, Estrella MM, Froissart M, et al. New Creatinine- and Cystatin C–Based Equations to Estimate GFR without Race. N. Engl. J. Med. 2021;385:1737–1749.

27. Elm E von, Altman DG, Egger M, Pocock SJ, Gøtzsche PC, Vandenbroucke JP, Initiative for the S. The Strengthening the Reporting of Observational Studies in Epidemiology (STROBE) statement: guidelines for reporting observational studies. J. Clin. Epidemiology. 2008;61:344– 349.

28. Zarin DA, Tse T, Williams RJ, Califf RM, Ide NC. The ClinicalTrials.gov Results Database — Update and Key Issues. N. Engl. J. Med. 2011;364:852–860.

29. American Community Survey 2019-2023 five-year estimates, tables B03002 (Hispanic or Latino Origin by Race), B01001B (Sex by Age, Black, or African American Alone), and B0100H (Sex by Age, White Alone, Not Hispanic or Latino) - Census Bureau Table [Internet]. [cited 2026 Sep 15];Available from: https://data.census.gov/table/ACSDT5Y2023.B03002

30. Akinbami LJ, Chen T-C, Davy O, Ogden CL, Fink S, Clark J, Riddles MK, Mohadjer LK. National Health and Nutrition Examination Survey, 2017-March 2020 Prepandemic File: Sample Design, Estimation, and Analytic Guidelines. Vital Heal. Stat. Ser 1, Programs Collect. Proced. 2022;1–36.

31. Saberi S, Wheeler M, Bragg-Gresham J, Hornsby W, Agarwal PP, Attili A, Concannon M, Dries AM, Shmargad Y, Salisbury H, et al. Effect of Moderate-Intensity Exercise Training on Peak Oxygen Consumption in Patients With Hypertrophic Cardiomyopathy: A Randomized Clinical Trial. Jama. 2017;317:1349.

32. Members WC, Ommen SR, Mital S, Burke MA, Day SM, Deswal A, Elliott P, Evanovich LL, Hung J, Joglar JA, et al. 2020 AHA/ACC Guideline for the Diagnosis and Treatment of Patients With Hypertrophic Cardiomyopathy A Report of the American College of Cardiology/American Heart Association Joint Committee on Clinical Practice Guidelines. J Am Coll Cardiol. 2020;76:e159–e240.

33. Axelsson A, Iversen K, Vejlstrup N, Ho C, Norsk J, Langhoff L, Ahtarovski K, Corell P, Havndrup O, Jensen M, et al. Efficacy and safety of the angiotensin II receptor blocker losartan for hypertrophic cardiomyopathy: the INHERIT randomised, double-blind, placebo-controlled trial. Lancet Diabetes Endocrinol. 2015;3:123–131.

34. Rader F, Oręziak A, Choudhury L, Saberi S, Fermin D, Wheeler MT, Abraham TP, Garcia-Pavia P, Zwas DR, Masri A, et al. Mavacamten Treatment for Symptomatic Obstructive Hypertrophic Cardiomyopathy Interim Results From the MAVA-LTE Study, EXPLORER-LTE Cohort. JACC: Hear. Fail. 2024;12:164–177.

35. Saberi S, Abraham TP, Choudhury L, Barriales-Villa R, Elliott PM, Nassif ME, Oreziak A, Owens AT, Tower-Rader A, Rader F, et al. Aficamten Treatment for Symptomatic Obstructive Hypertrophic Cardiomyopathy 48-Week Results From FOREST-HCM. JACC: Hear. Fail. 2025;13:102496.

36. Arias E, Schauman WS, Eschbach K, Sorlie PD, Backlund E. The validity of race and Hispanic origin reporting on death certificates in the United States. Vital Heal. Stat. Ser. 2, Data Eval. methods Res. 2008;1–23.

37. Ingram DD, Parker JD, Schenker N, Weed JA, Hamilton B, Arias E, Madans JH. United States Census 2000 population with bridged race categories. Vital Heal. Stat. Ser. 2, Data Eval. methods Res. 2003;1–55.

38. Reza N, Butzner M, Batra K, Amos Q, Buikema A, Shreay S, Owens A. Impact of Sociodemographic Characteristics on Outcomes in Obstructive Hypertrophic Cardiomyopathy. J. Am. Hear. Assoc.: Cardiovasc. Cerebrovasc. Dis. 2026;15:e044294.

39. Hafeez N, Claggett BL, Owens AT, Helms AS, Saberi S, Lampert R, Stendahl JC, Ashley EA, Parikh VN, Lakdawala NK, et al. Social Determinants of Health and Clinical Outcomes in Hypertrophic Cardiomyopathy. JAMA Cardiol. 2026;11:165–174.

40. Eberly LA, Day SM, Ashley EA, Jacoby DL, Jefferies JL, Colan SD, Rossano JW, Semsarian C, Pereira AC, Olivotto I, et al. Association of Race With Disease Expression and Clinical Outcomes Among Patients With Hypertrophic Cardiomyopathy. JAMA Cardiol. 2020;5:83–91.

41. Galsky MD, Stensland KD, McBride RB, Latif A, Moshier E, Oh WK, Wisnivesky J. Geographic Accessibility to Clinical Trials for Advanced Cancer in the United States. JAMA Intern. Med. 2015;175:293–295.

42. Hantel A, Kohlschmidt J, Eisfeld A-K, Stock W, Jacobson S, Mandrekar S, Larson RA, Stone RM, Lathan CS, DeAngelo DJ, et al. Inequities in Alliance Acute Leukemia Clinical Trial and Biobank Participation: Defining Targets for Intervention. J. Clin. Oncol. 2022;40:3709–3718.

43. Hawk ET, Habermann EB, Ford JG, Wenzel JA, Brahmer JR, Chen MS, Jones LA, Hurd TC, Rogers LM, Nguyen LH, et al. Five National *Cancer* Institute–designated cancer centers’ data collection on racial/ethnic minority participation in therapeutic trials. Cancer. 2014;120:1113–1121.

44. Tai CG, Hiatt RA. The Population Burden of Cancer: Research Driven by the Catchment Area of a Cancer Center. Epidemiologic Rev. 2017;39:108–122.

45. Parker JD, Talih M, Malec DJ, Beresovsky V, Carroll M, Gonzalez JF, Hamilton BE, Ingram DD, Kochanek K, McCarty F, et al. National Center for Health Statistics Data Presentation Standards for Proportions. Vital Heal. Stat. Ser. 2, Data Eval. methods Res. 2017;1–22.

46. Johnson CL, Paulose-Ram R, Ogden CL, Carroll MD, Kruszon-Moran D, Dohrmann SM, Curtin LR. National health and nutrition examination survey: analytic guidelines, 1999-2010. Vital Heal. Stat. Ser. 2, Data Eval. methods Res. 2013;1–24.

47. Paule RC, Mandel J. Consensus Values and Weighting Factors. J. Res. Natl. Bur. Stand. 1982;87:377.

48. Higgins JPT, Thompson SG, Deeks JJ, Altman DG. Measuring inconsistency in meta-analyses. BMJ. 2003;327:557–560.

49. Higgins JPT, Thompson SG. Quantifying heterogeneity in a meta-analysis. Stat. Med. 2002;21:1539–1558.

50. Newcombe RG. Two-sided confidence intervals for the single proportion: comparison of seven methods. Stat. Med. 1998;17:857–872.

51. Barendregt JJ, Doi SA, Lee YY, Norman RE, Vos T. Meta-analysis of prevalence. J. Epidemiology Community Heal. 2013;67:974.

52. Dias KA, Link MS, Levine BD. Exercise Training for Patients With Hypertrophic Cardiomyopathy JACC Review Topic of the Week. J Am Coll Cardiol. 2018;72:1157–1165.

53. Ho CY, Mealiffe ME, Bach RG, Bhattacharya M, Choudhury L, Edelberg JM, Hegde SM, Jacoby D, Lakdawala NK, Lester SJ, et al. Evaluation of Mavacamten in Symptomatic Patients With Nonobstructive Hypertrophic Cardiomyopathy. J. Am. Coll. Cardiol. 2020;75:2649–2660.

54. Maron MS, Masri A, Choudhury L, Olivotto I, Saberi S, Wang A, Garcia-Pavia P, Lakdawala NK, Nagueh SF, Rader F, et al. Phase 2 Study of Aficamten in Patients With Obstructive Hypertrophic Cardiomyopathy. J. Am. Coll. Cardiol. 2023;81:34–45.

55. Masri A, Lester SJ, Stendahl JC, Hegde SM, Sehnert AJ, Balaratnam G, Shah A, Fox S, Wang A. Long-Term Safety and Efficacy of Mavacamten in Symptomatic Obstructive Hypertrophic Cardiomyopathy: Interim Results of the PIONEER-OLE Study. J. Am. Hear. Assoc.: Cardiovasc. Cerebrovasc. Dis. 2024;13:e030607.

56. Desai MY, Owens A, Geske JB, Wolski K, Naidu SS, Smedira NG, Cremer PC, Schaff H, McErlean E, Sewell C, et al. Myosin Inhibition in Patients With Obstructive Hypertrophic Cardiomyopathy Referred for Septal Reduction Therapy. J. Am. Coll. Cardiol. 2022;80:95–108.

57. Batchelor WB, Damluji AA, Yong C, Fiuzat M, Barnett SD, Kandzari DE, Sherwood MW, Epps KC, Tehrani BN, Allocco DJ, et al. Does study subject diversity influence cardiology research site performance?: Insights from 2 U.S. National Coronary Stent Registries. Am. Hear. J. 2021;236:37–48.

58. Clark LT, Watkins L, Piña IL, Elmer M, Akinboboye O, Gorham M, Jamerson B, McCullough C, Pierre C, Polis AB, et al. Increasing Diversity in Clinical Trials: Overcoming Critical Barriers. Curr. Probl. Cardiol. 2019;44:148–172.

59. Sorensen LL, Pinheiro A, Dimaano VL, Pozios I, Nowbar A, Liu H, Luo H-C, Lin X, Olsen NT, Hansen TF, et al. Comparison of Clinical Features in Blacks Versus Whites With Hypertrophic Cardiomyopathy. Am. J. Cardiol. 2016;117:1815–1820.

60. Markham DW, Dries DL, King LP, Leonard D, Yancy CW, Peshock RM, Willett D, Cooper RS, Drazner MH. Blacks and whites have a similar prevalence of reduced left ventricular ejection fraction in the general population: The Dallas Heart Study (DHS). Am. Hear. J. 2008;155:876–882.

61. Yao K, Bai Y, Kang X, Zhang Y, Qiao J. Trends in the prevalence of apparent treatment-resistant hypertension among U.S. adults, 1999–2018. Am. J. Prev. Cardiol. 2025;24:101360.

62. Bierer BE, White SA, Gelinas L, Strauss DH. Fair payment and just benefits to enhance diversity in clinical research. J. Clin. Transl. Sci. 2021;5:e159.

63. Williams CP, Geiger AM, Norton WE, Moor JS de, Everson NS. Influence of Cost-Related Considerations on Clinical Trial Participation: Results from the 2020 Health Information National Trends Survey (HINTS). J. Gen. Intern. Med. 2023;38:1200–1206.

